# Visualizing and Assessing US County-Level COVID19 Vulnerability

**DOI:** 10.1101/2020.07.30.20164608

**Authors:** Gina Cahill, Carleigh Kutac, Nicholas L. Rider

## Abstract

**Objective:** Like most of the world, the United States’ public health and economy are impacted by the COVID19 pandemic. However, discrete pandemic effects may not be fully realized on the macro-scale. With this perspective, our goal is to visualize spread of the pandemic and measure county-level features which may portend vulnerability.

**Materials and Methods:** We accessed the New York Times GitHub repository COVID19 data and 2018 US Census data for all US Counties. The disparate datasets were merged and filtered to allow for visualization and assessments about case fatality rate (CFR%) and associated demographic, ethnic and economic features.

**Results:** Our results suggest that county-level COVID19 fatality rates are related to advanced population age (p <0.001) and less diversity as evidenced by higher proportion of Caucasians in High CFR% counties (p < 0.001). Also, lower CFR% counties had a greater proportion of the population reporting has having 2 or more races (p <0.001). We noted no significant differences between High and Low CFR% counties with respect to mean income or poverty rate.

**Conclusions:** Unique COVID19 impacts are realized at the county level. Use of public datasets, data science skills and information visualization can yield helpful insights to drive understanding about community-level vulnerability.

## Introduction

In December 2019, an unknown form of pneumonia was identified in Wuhan Province China which ultimately heralded emergence of the COVID19 pandemic. Since that time, spread of SARS-CoV-2 resulted in over 14 million individuals infected and over 600,000 deaths worldwide[1, 2]. At present, COVID19 has spread to every continent on Earth except for Antarctica.[2, 3] In the United States(US), the first confirmed COVID19 case was reported by a team in Snohomish County Washington.[4] In the ensuing 7 months, the US reported over 3.8 million infections and over 140,000 deaths.[2] Given the totality of health and economic devastation levied by COVID19, tremendous energy has been invested towards understanding features which could predict individual and community impact.[5-10]

Understanding community vulnerability with COVID19 is important and may enable pre-emptive and ongoing interventions to stem public health and economic crises.[11-13] However the explosion of data about COVID19 has led to much debate and skepticism around best measures for analysis and assessment of impact.[14] Now that widespread disease reporting is in place, case fatality rate (CFR%) can reasonably be used.[15-17] With widespread US testing and reporting of COVID19 cases, we used assessment of CFR% across US counties coupled with US Census data to visualize and evaluate regional differences in COVID impact. Our analysis was focused on 4 months of pandemic activity (March – June 2020) for US Counties. Here we show pandemic spread via CFR% visualized across US counties stratified by both CFR% and case burden. Additionally, we show relevant Census-mined data attributes associated with high and low CFR% counties.

## Materials and Methods

We extracted US county data about the COVID19 pandemic from the New York Times Github repository (https://github.com/nytimes/covid-19-data) and US Census data (2018) about county-level demographics, economic factors and ethnicities (https://www.census.gov). Data was extracted on July 6, 2020. Using JupyterLab through the Anaconda distribution (v. 2020.02) Python version 3.8.3 with Pandas (version 1.0.5) we merged and filtered the final dataset for analysis. Our code is available here (https://github.com/nlrider/COVID-Public-Health-Data). Unique counties with a CFR% between 0.1 and 100 and a Federal Information Processing Standard (fips) code were analyzed for the last date of each month studied (March – June 2020). Case fatality rate % was calculated by taking the ratio of absolute deaths per county at the given time by absolute total county cases and multiplying by 100.

From the list of unique counties with a CFR% between 0.1-100, we assessed median case count per month per county. We then visualized both the total number of counties available at end of each month and also counties above the median by relative scale CFR% (Figure 1: A-D). County level data visualization was done with plotly (v. 4.8.2) in JupyterLab for available counties by fips codes at monthly time intervals reported on the last date of each month.

**Figure 1A-D:**
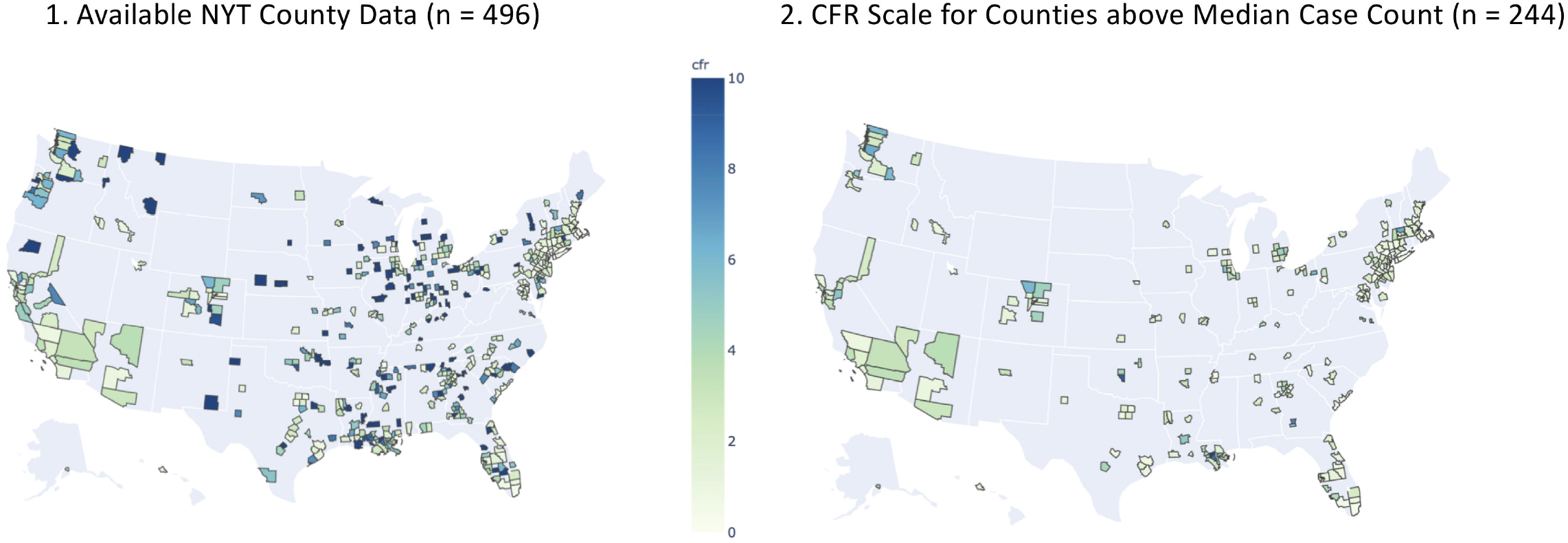
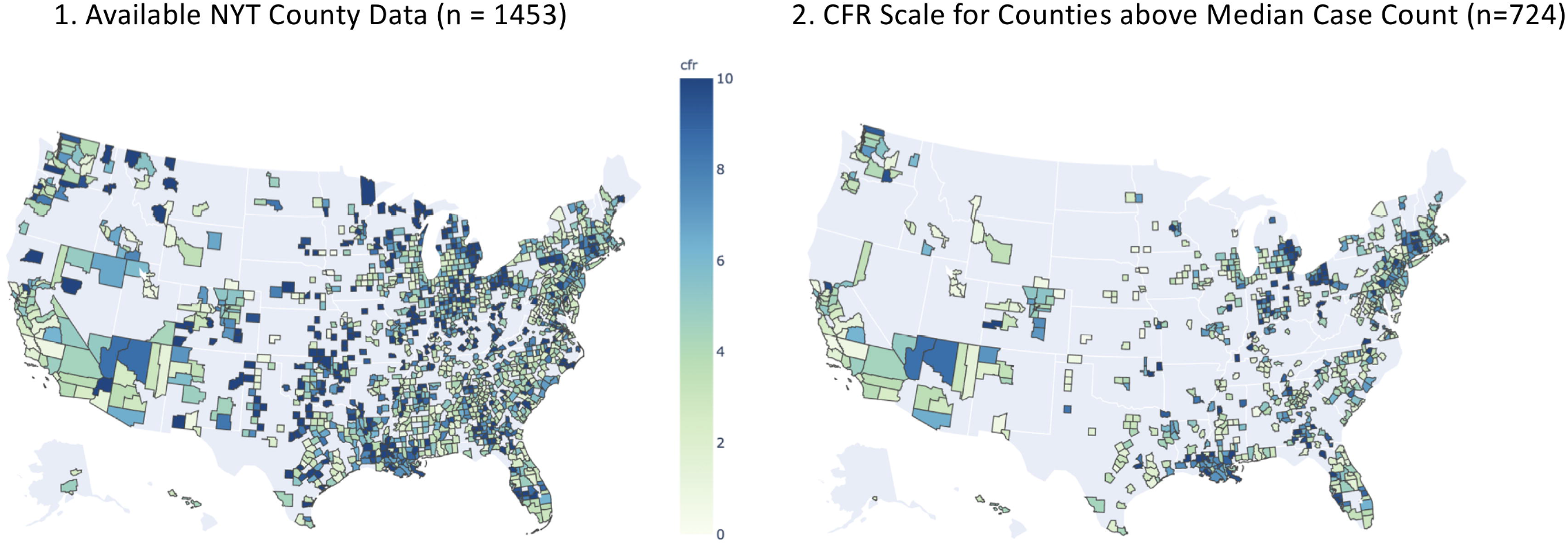
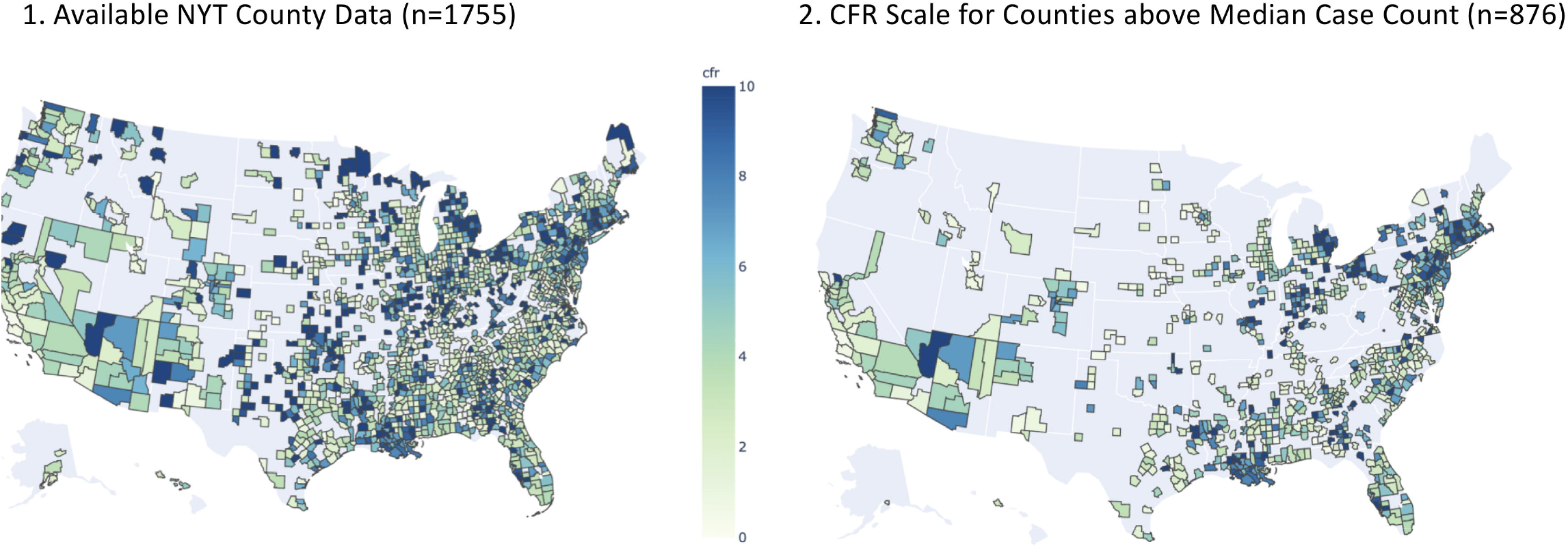
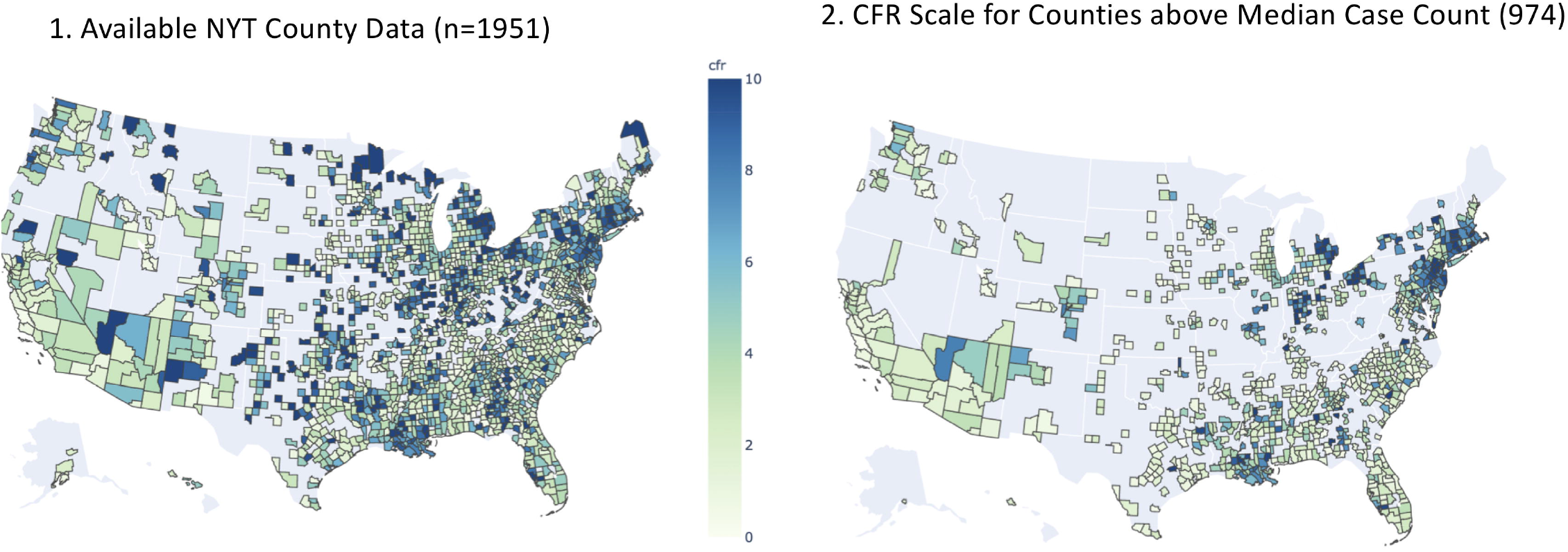
Each panel shows US Counties visualized by relative scale CFR% for the specified month. Left side panels display all available counties within the CFR% range (0.1-99.9) and right-side panels show CFR% for counties with case counts about the National median. Horizontal bar-scale shows relative CFR% as plotted from 0 - 10 relative. County numbers shown above each map.

Descriptive statistics for each month time capsule was performed in JupyterLab using Pandas (Supplemental Tables 1-4). High CFR counties as of June 30, 2020 were defined by a county above the median case count and with a CFR% at or above the upper quartile (>6.25%). Low CFR counties were also taken from all counties with a case count above the median but with a CFR% falling below the upper quartile (6.25% or lower). Subsequent significance testing for county data as of June 30, 2020 was performed with STATA version 12.1 Welch’s t-tests were used to determine whether or not there was a significant difference in the mean values of variables of interest (NYT COVID and US Census Features) between high and low CFR counties, as this method of testing accounts for unequal variances between the two groups (Table 2). Rank of US states by number of high or low CFR counties as of June 30, 2020 were plotted and sorted using Tableau (v.2020.2; Figure 2 and 3).

**Table 2:** County-level Features Comparing High and Low CFR Counties.

| County Feature | CFR Class | Mean (STD) | p-value |
| --- | --- | --- | --- |
| Total Population | Low | 159,128(438,503) | 0.007 |
|  | High | 117,591(226,235) |  |
| Population Below Poverty Line | Low | 22,539(66,667) | 0.003 |
|  | High | 14,570(29,438) |  |
| % Population Below Poverty Line | Low | 15.9(0.17) | 0.222 |
|  | High | 15.6(0.30) |  |
| Total Population > 16 Years of Age | Low | 129,470(354,037) | 0.01 |
|  | High | 97,621(186,687) |  |
| % Population > 16 Years of Age | Low | 82.8(5.9) | 0.001 |
|  | High | 83.9(5.7) |  |
| Unemployment Rate | Low | 6.1(2.5) | 0.52 |
|  | High | 5.9(2.6) |  |
| Total Population > 62 Years of Age | Low | 29,365(73.066) | 0.1 |
|  | High | 24,800(45,203) |  |
| % Population > 62 Years of Age | Low | 21.4(4.9) | <0.001 |
|  | High | 23.5(4.5) |  |
| Median Income (USD) | Low | 53,006(14,964) | 0.97 |
|  | High | 52,982(15,809) |  |
| Population >25 Years of Age and HS Graduate | Low | 28,585(66,645) | 0.07 |
|  | High | 24,008(40,751) |  |
| % Population >25 Years of Age and HS Graduate | Low | 23.3(6.2) | <0.001 |
|  | High | 25.2(5.7) |  |
| Population with College Degree | Low | 21,505(65,331) | 0.06 |
|  | High | 16,850(38,256) |  |
| % Population with College Degree | Low | 10.2(4.1) | 0.95 |
|  | High | 10.2(4.2) |  |
| Total Caucasian Population | Low | 116,080(271,635) | 0.03 |
|  | High | 93,703(16,829) |  |
| % Population Caucasian | Low | 81.5(17.6) | <0.001 |
|  | High | 84.8(17.9) |  |
| Total Black Population | Low | 21,520(71,286) | 0.01 |
|  | High | 14,496(45,623) |  |
| % Population Black | Low | 13.7(17.6) | 0.1 |
|  | High | 12.1(18.2) |  |
| Total American Indian Population | Low | 1322(4888) | <0.001 |
|  | High | 707(2637) |  |
| % Population American Indian | Low | 1.4(5.9) | 0.8 |
|  | High | 1.4(5.2) |  |
| Total Asian Population | Low | 9430(56,395) | 0.01 |
|  | High | 5204(18,778) |  |
| % Population Asian | Low | 1.9(3.2) | 0.01 |
|  | High | 1.5(2.5) |  |
| Total Hawian Native Population | Low | 352(2915) | <0.001 |
|  | High | 47(113) |  |
| % Hawian Native | Low | 0.1(0.4) | <0.001 |
|  | High | 0.04(0.09) |  |
| Population with 2 or More Races | Low | 5563(18,319) | <0.001 |
|  | High | 3203(6689) |  |
| % Population with 2 or More Races | Low | 2.6(1.9) | <0.001 |
|  | High | 2.3(1.5) |  |
| CFR = Case Fatality Rate |  |  |  |
| p-value calculted with Welch's t-test for unequal variances |  |  |  |

**Figure 2:**
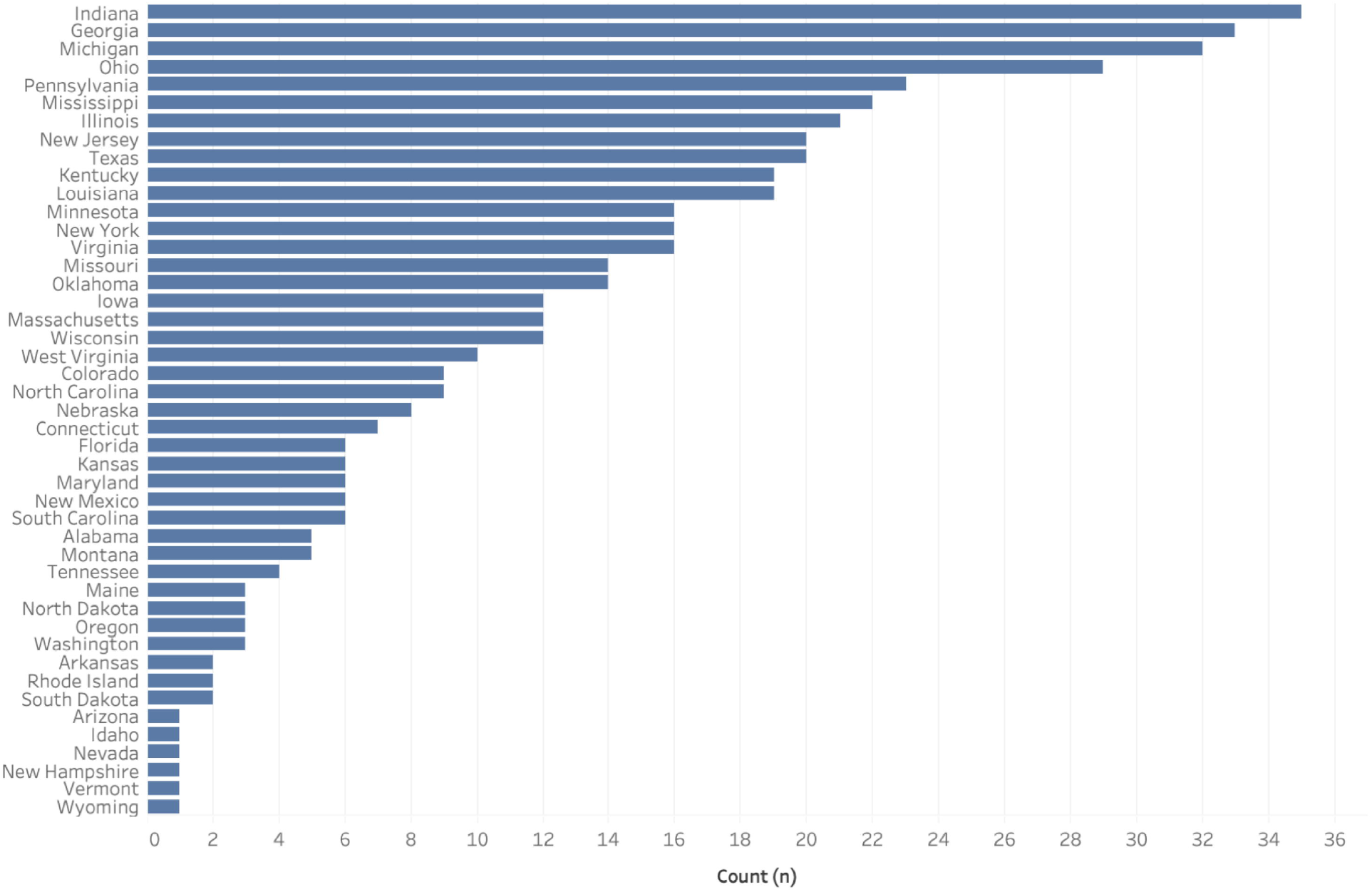
Horizontal bar chart depiction of states and corresponding county counts within the highest US National quartile (i.e. CFR% > 6.25). States sorted in descending order.

**Figure 3:**
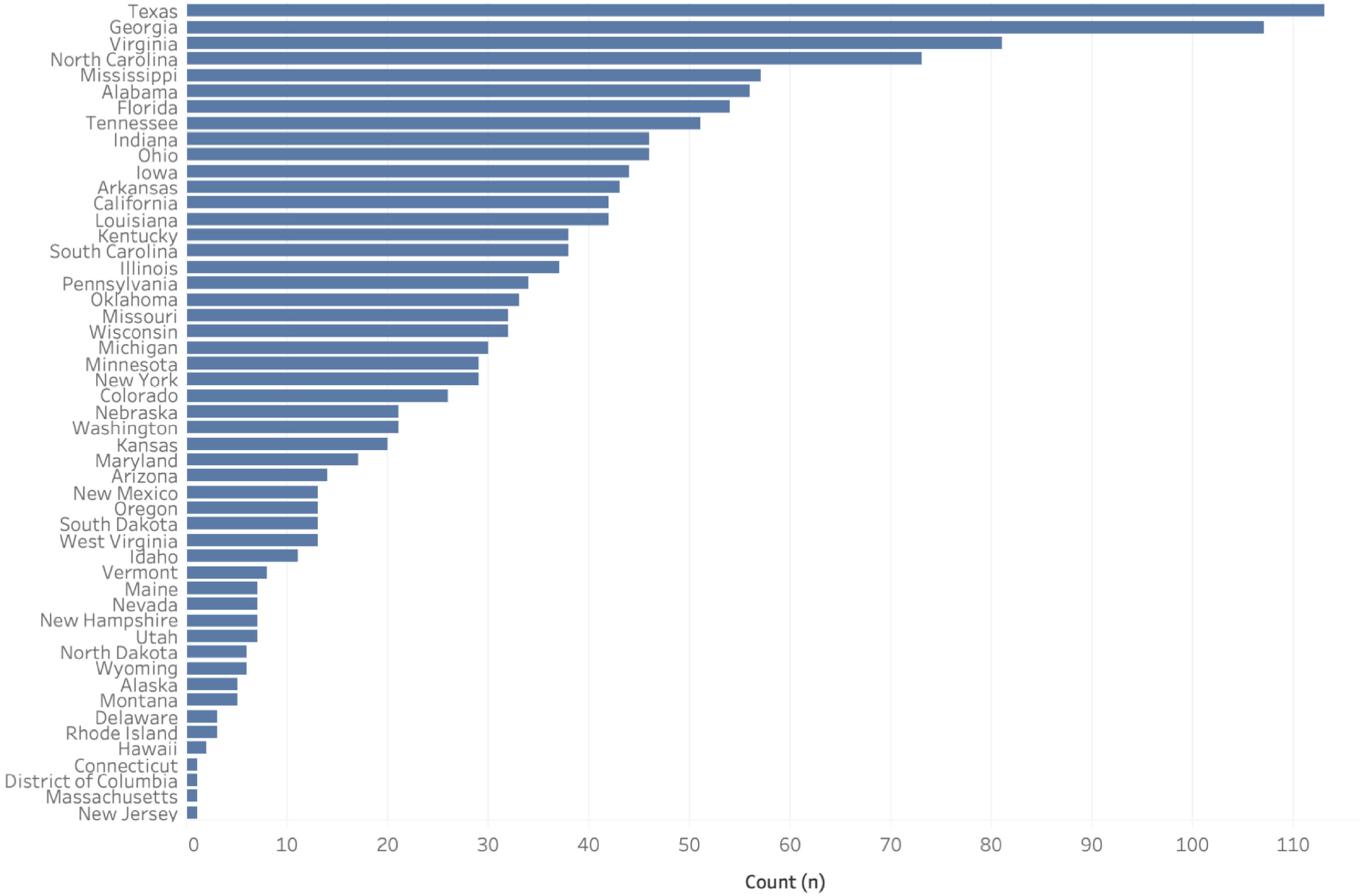
Horizontal bar chart depiction of states and corresponding county counts within the lower 3 US National quartiles (i.e. CFR% ≤ 6.25). States sorted in descending order.

## Results

Data extraction from the New York Times COVID19 GitHub repository (https://github.com/nytimes/covid-19-data) revealed 3060 unique fips reflecting the same number of counties and associated COVID data. Filtering data to include counties with a CFR% between 0.1%-100% reduced this number of unique counties to 1996. This number of counties was the total available for our 4-month analysis, however pandemic data available at each monthly time interval varied as shown in Table 1. For each month interval assessed, an increase in the number of counties available for analysis is noted in alignment with the expansion of the pandemic. Similarly, the median per county case count increased and was paralleled by deaths across the 4 months assessed (244 to 974 cases/county). However, slope of the increase in case counts and deaths reported was not identical as noted by the gradual trend down in CFR% from end of March to end of June (7.1% to 4.6%) as seen in Table 1.

**Table 1:** Aggregate County-level COVID19 Statistics by Month Studied.

| **Table 1: Aggregate County-level COVID19 Statistics by Month Studied** |  |  |  |  |
| --- | --- | --- | --- | --- |
|  | **March** | **April** | **May** | **June** |
| **County Count (n)** | 496 | 1453 | 1755 | 1951 |
| **Mean Case Count per County (n)** | 339 | 722 | 1007 | 1337 |
| **Median Case Count per County (n)** | 55 | 88 | 144 | 228 |
| **Mean Death Count per County (n)** | 8 | 43 | 59 | 65 |
| **Mean County CFR(%)** | 7.1 | 6.3 | 5.3 | 4.6 |
| **Counties above Median Case Count/Month (n)** | 244 | 724 | 876 | 974 |

Progression of the pandemic by CFR% at the last date of each month is shown in Figure 1 with the associated high-case counties plotted according to CFR%. The left-side map in each panel shows total cases for counties that met our criteria by CFR%; whereas, the right-side map shows CFR% for counties with case counts above the US median. The CFR% scale is arbitrary and only shows relative differences.

State level county assessments are shown in Figure 2 and 3. Figure 2 shows the county counts for each state sorted in descending order within the high CFR% category (CFR% > 6.25; upper quartile). Figure 3 shows state-level county counts of those within the bottom 3 quartiles (CFR% ≤ 6.25). Figure 4 shows all available state data without lower or upper boundaries for case count as of July 6, 2020.

**Figure 4:**
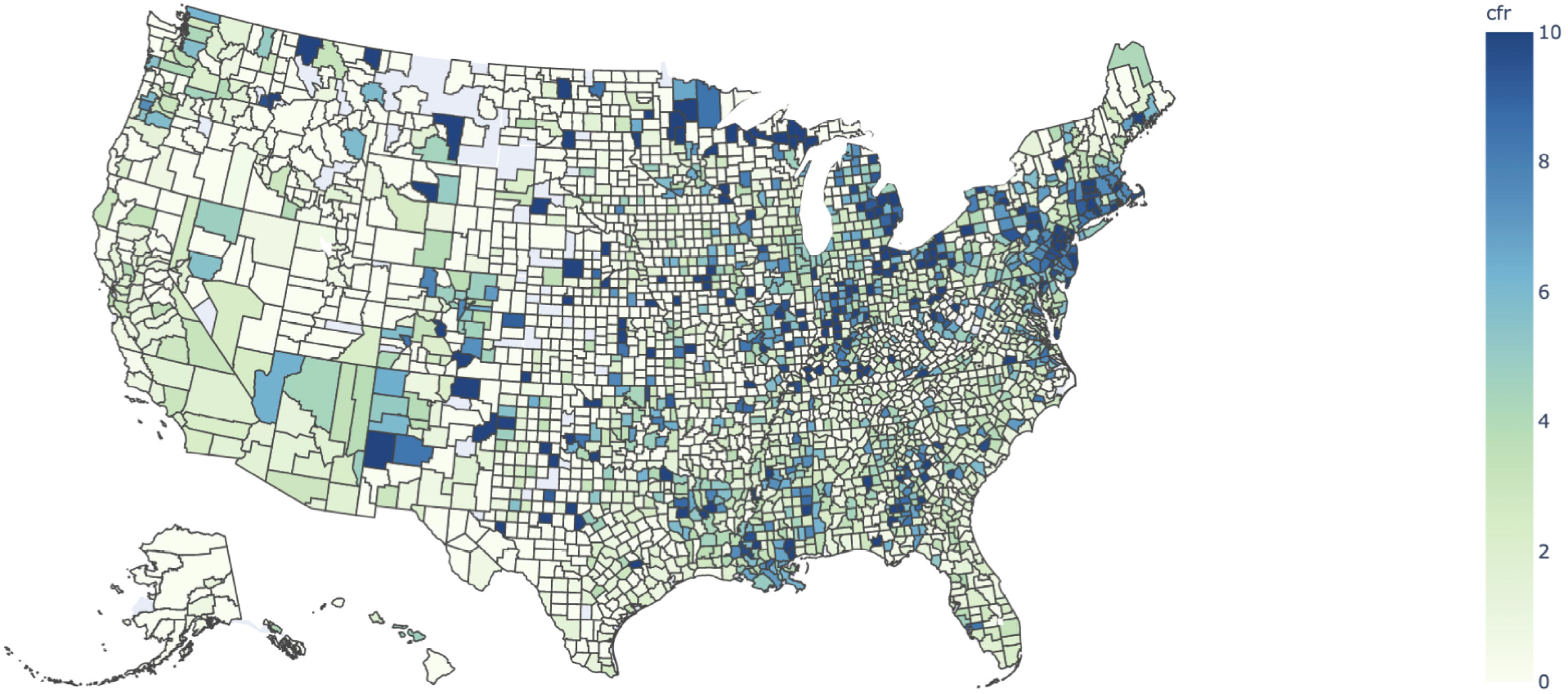
Plot of all available US County CFR% data (n = 3060) without case count filter. Color range in relative scale 0 – 10 as of July 6, 2020.

Descriptive aggregate COVID19 statistics per month studied are shown in Table 1. Table 2 shows significance testing across socioeconomic, demographic and ethnic county features in comparing top quartile counties with those in the bottom 3 quartiles. From this analysis we found that significant, normalized differences between High CFR% and Low CFR% counties included more of the following in High CFR% counties: % of population over 16 years of age, % of population above 62 years of age, % of population over 25 years of age with a high school degree, % of population reporting as Caucasian. We also noted significantly higher numbers of the following in Low CFR% counties: total and % of population reporting as Asian, total and % of population reporting as Pacific Island Native/Hawaiian as well as total and % of counties reporting 2 or more races.

## Discussion

Spread of COVID19 across the US has been rapid and hot spots such as New York City, Seattle Washington, New Orleans Louisiana and Los Angeles California emerged early and can be seen in our mapped data (Figure 1: A1&2).[18, 19] We note persistence of “hot spot” High CFR% counties within the Northeast, the Gulf Coast region, the Upper Midwest and Desert Southwest, Southern California and the Pacific Northwest (Figure 1: Progression from A to D). Notably, the only means by which a county’s CFR% would drop over time is to have more people contract COVID19 and survive or to have the ongoing proportion of death/confirmed cases diminish with time. We do see such a dynamic occur via the 4-month time course (Table 1). Also, the reduction of mean county CFR% shown in Table 1 from March to June reflects a sharp rise in mean case count over the timespan.

Interestingly, our visualizations show that High CFR% US counties are not isolated to urban regions (Figure 1:A-D, Figure 3). Rather, wide distribution of high CFR% counties suggests that unique demographic, geographic or other features may affect county-level vulnerability. Our CFR% mapping, not surprisingly aligns with other group’s work related to community risk.[20, 21] Additionally, an individual state may have a significant number of both High and Low CFR% Counties as shown in Figures 2 & 3. If we focus on the top 10 states with most High CFR% counties, we see that they represent the Midwest, South East, South, and Northeast regions. Top 10 states for Low CFR% county counts also overlap with the High CFR% group in that Texas, Georgia, Mississippi, Indiana and Ohio are present in both groups (Figure 2 & 3). This underscores the notion that community vulnerability may be intrinsic and likely independent of climate, local governmental policy or even population density. It is also important to note that a priori county-level risk and actual CFR% are not uniformly aligned.

Our assessment of county-level features which may contribute to higher or lower CFR% included Census information taken from demographic, economic and ethnicity tables. From these, our data suggest that a more diverse and younger population is better able to weather COVID19 (Table 2). This census-based analysis on an unbalanced set of counties with High CFR% (highest quartile) with a CFR% > 6.25 to that of the bottom 3 quartiles, shows that predominantly Caucasian counties (% Caucasian) were found in greater numbers within the High CFR% group. Conversely, counties with proportionally more Asians and Pacific Islander/Hawaiian Natives were found in the Low CFR% group. We also note that counties reporting as having a higher proportion of the population representing 2 or more races, were more likely to be in the Low CFR% county group. Importantly, no differences were observed between High and Low CFR% groups in their levels of unemployment, median income or proportion of population below the poverty line. Also, education level did not seem to differ in that college degree holders were in similar numbers across both groups. While there were more high school diplomates over age 25 years in the High CFR%, this may relate to risk associated with age.

The COVID19 pandemic remains dynamic without clear evidence for which communities or individuals may fare worse from the outset. Factors such as population health features, number of long-term care facilities, prisons or other workplaces known to drive outbreaks are important and were not addressed here.[22-24] Also, analysis of a comprehensive public health dataset is important for teasing out community-level risks. However, no single or amalgamated resource retrospectively analyzed will capture all needed features for complete risk assessment. Lastly, some of our results are in contrast to previously reported findings which show important regional ethnic risk factors for outcomes in COVID19.[25] Such differences may relate to intrinsic reporting bias with US Census data as noted previously.[26]

In summary, we present an analysis of county-level CFR% via merged COVID19 and US Census data. Our findings suggest that younger and more diverse counties fared better over the 4 months studied for the US COVID19 experience. We also find that community risk level must be assessed at granularity level below that of the state or region as scrutiny of specific population-level features may yield greater insights for enabling population resiliency. Implementation of policies and systems which foster prospective quality assessments and enable rigorous analysis are needed for the US Healthcare System and will likely need to be implemented at that level of detail.

## Data Availability

Data outside of supplementary material will be made available upon reasonable request.

## Acknowledgements

We wish to thank the team and investigators at the New York Times for making COVID19 project and associated GitHub repository data publicly available here - https://github.com/nytimes/covid-19-data

